# Correlation between life satisfaction and symptoms of attention deficit hyperactivity disorder (ADHD) in dental students: The mediation of resilience

**DOI:** 10.1101/2023.05.15.23290010

**Authors:** Yuwei Sun, Lei Miao, Siying Chen, Zhenya Piao, Chi Tong

## Abstract

The aim of the study was to examine the morbidity of Attention Deficit Hyperactivity Disorder (ADHD) symptoms in dental students, explore the correlation between life satisfaction and ADHD symptoms. It also discusses whether resilience mediates the correlation between ADHD symptoms and life satisfaction. Self-report questionnaires consist of the Wender Utah Rating Scale (WURS), the Adult ADHD Self-Report Scale (ASRS), the Satisfaction With Life Scale (SWLS), the Conner-Davidson Resilience Scale (CD-RISC), and sociodemographic characteristics. The analyses with Hierarchical linear regression were operated to investigate the effect of ADHD symptoms on life satisfaction. The study used resampling and asymptotic strategies to discuss the mediation of resilience. 291 dental students became final objects. Twenty students (6.87%) may have ADHD. There were differences in ADHD prevalence among objects of diverse ages and with varying levels of paternal education. The ADHD asymptomatic group had distinctly higher resilience and life satisfaction degrees than the symptomatic group. Inattention and hyperactivity were both correlated negatively with life satisfaction and resilience. Life satisfaction was observably positively associated with resilience. Resilience serves as a mediating role between life satisfaction and the two symptoms of ADHD. Detecting ADHD symptoms earlier is helping improve mental health of future dentists. Resilience intervention programs can enhance life satisfaction of dental students.

## Introduction

A global study pointed out that mental disorders’ burden increased from 1990 to 2019 [1]. Attention deficit hyperactivity disorder (ADHD) is a sort of the most familiar mental disorders affecting nonage and adults, which leads to an enormous economic burden worldwide [2]. The main features of ADHD incorporate inattention, hyperactivity, and impulsivity [3]. With the progress of many cohort studies, more and more scholars have recognized that ADHD symptoms usually appear in childhood, and a significant proportion persists into adolescence or adulthood. The prevalence in children is about 5% and is twice as high in boys as in girls [4]. The World Mental Health Survey published by the World Health Organization(WHO) points out that the prevalence of adults is about 2.8% [5]. Chronic symptoms of ADHD have invariably been shown to be related to impairment in many fields of life, like occupational achievement, social correlations, substance use, academic record, and other psychiatric comorbidities [6–10]. Nevertheless, few studies have statistically analyzed the prevalence of ADHD in dental students by searching mainstream databases such as PubMed and Web of Science. With the continuous development of Psychiatry, Psychology, and Education, more and more people began concentrating on ADHD in college students. However, studies on medical students have mainly focused on the adverse effects, including suicidal behaviour and Internet addiction, accompanying ADHD[11–13]. At the same time, the study on the association between ADHD and positive outcomes, including quality of life and life satisfaction, is minimal. In scientific efforts, clinical practice, and teaching practice, the dimensional approach of ADHD can allow people to obtain more information than the traditional classification method [14]. A study of middle-aged and older Australians showed that inattention was more relevant than hyperactivity in ADHD symptoms in social, academic, occupational functioning, and psychological outcomes, which include life satisfaction [15]. Until now, however, no study has examined this issue among dental students. Although ADHD symptoms may generate impairment in some related functions, they do not significantly impact some people’s lives. According to our conjecture, this group may have positive psychological resources to cope with adversity better. Since positive psychology was proposed to prevent and treat mental health problems and improve people’s overall subjective well-being, positive psychology has been increasingly applied to many people, including children, doctors, and soldiers [16–18]. As one of the positive psychological resources, resilience can reduce the negative impact of psychiatric symptoms in ADHD patients and improve their life satisfaction [19,20]. Generally speaking, resilience is how people understand, manage, and control themselves after a significant stressful or traumatic event [21]. It can be inferred from the transaction model of coping and stress[22] that resilience, as a psychological resource, can quadratically affect the evaluation process of stressors and thus moderate the relationship between stressors and their outcomes. Still, there are few studies related to the potential effects of resilience in the correlation between life satisfaction and ADHD symptoms. Given the increasing socioeconomic burden of ADHD, existing studies on ADHD symptoms are significantly underrepresented in dental students. We conducted the research to 1) analyze the morbidity of ADHD symptoms in dental students of China; 2) explore the relationship between life satisfaction and two segments of ADHD symptoms among dental students; 3) explore the mediation between life satisfaction and ADHD symptoms.

## Materials and Methods

### Subjects and Study design

The research was a cross-sectional investigation. It was carried out in May 2023 at a medical university in China. “Wenjuanxing” (www.wjx.cn), a famous professional online survey platform, was used to conduct questionnaires and collect data. During or after data capture, authors can access information that identifies individual participants. A stratified cluster sampling method was operated to recruit dental students based on academic years. All students participated voluntarily and were informed of the study’s purpose before answering the questionnaire. At the beginning of the questionnaire, we designed an informed consent link. The subjects who answered the questionnaire were considered to have signed a written informed consent agreement before answering the questionnaire. Medical Ethics Committee of China Medical University authorized the study.

### Assessment of ADHD symptoms in adults

The study adopted the Chinese edition of the Adult ADHD Self-Report Scale(ASRS) to measure adult ADHD symptoms in subjects. [23]. This scale has two subdivisions named inattention and hyperactivity. Both have nine entries. (e.g., “How often do you fidget or squirm with your hands or feet when you have to sit down for a long time?” and “How often do you misplace or have difficulty finding things at home or work?”.) Each article is ranked on a Likert scale with five points from zero (“never”) to four (“very often”). The respondents were requested to score each article owing to their affairs in the last six months Subjects who scored more than 24 would have ADHD in all probability, those who scored 17-23 were sorted into possibly having ADHD, and those who scored 0-16 were almost not likely to have ADHD [24]. Owing to previous studies, the Chinese edition of ASRS has decent credibility and validity. [23–25]. Cronbach’s alpha for the inattention segment was 0.78 and the hyperactivity dimension was 0.85.

### Assessment of ADHD symptoms in nonage

The Wender Utah Rating Scale (WURS) was operated to measure ADHD symptoms in nonage among the objects. The scale with twenty-five items was used primarily to review individual ADHD symptoms during adolescence. Sample items include “When I was a child, I had trouble seeing things from someone else’s point of view” and “As a child, I was sad or blue, depressed, unhappy”. Each article is ranked on a Likert scale with five points from zero (“not at all or very slightly”) to four (“very much”). The researchers used an aggregate score of 46 as the cut-off point for childhood ADHD [26]. Scholars believe WURS has decent credibility and validity. Cronbach’s alpha in the research was 0.94.

### Classification criteria of ADHD symptomatic set and asymptomatic set

In clinical practice, only individuals with both overt childhood ADHD symptoms and current ADHD symptoms are diagnosed as adult ADHD. Therefore, subjects with WURS scores ≥46 and at a minimum one section of ASRS scores ≥17 were divided into a symptomatic set in this study, and the remaining were classified as asymptomatic.

### Measurement of life satisfaction

The Satisfaction With Life Scale (SWLS) estimates the worldwide aspect of life satisfaction [27]. The scale owns five items (e.g., “So far, I have gotten the influential things I want in life” and “I am satisfied with my life”). Each article is responded to on a Likert scale with seven points from one (“strongly disagree”) to seven (“strongly agree”). The lower the ranking scored, the less satisfied the subjects were with their lives. Past researches have demonstrated convincing credibility and predictive validity among diverse age groups [27–29]. Cronbach’s alpha for the scale was 0.91

### Assessment of resilience

The Connor–Davidson Resilience Scale (CD-RISC) was developed to estimate the capacity to deal with pressure and adverse circumstances [30]. CD-RISC, possessing twenty-five items, has a higher psychometric property rating than most scales that measure resilience [31]. Sample items include “As things look hopeless, I do not give up condition” and “Past success gives me confidence for new challenges.” Each article is ranked on a Likert scale with five points from zero (“not true at all”) to four (“accurate nearly all the time”). The subjects with lower scores have a lower standard of resilience. In China, owing to the positive validity and credibility of CD-RISC, it has been made widely use of [30,31]. Cronbach’s alpha for CD-RISC was 0.94.

### Demographic characteristics

Demographic information for the study incorporated gender, age, race, place of residence, academic year, whether the only child and parental educational level.

### Statistical analysis

SPSS 26.0 was operated to analyze all the data. The statistical examinations we used were completely two-sided, and results with P < 0.05 were marked as remarkable. The study used T-tests and chi-squared tests to examine the distinctions in psychological and categorical variables between ADHD symptomatic set and the asymptomatic set. Pearson’s correlation analysis was conducted to check associations among life satisfaction, resilience, inattention and hyperactivity. The researchers investigate the influence of independent variables on life satisfaction using hierarchical regression analysis. F, R2, R2-changes (ΔR2) and Standardized estimates (β) were supplied for each step. Resampling and asymptotic strategies, which were developed by Hayes and Preacher [29], were operated to check the mediation of resilience (a*b) on the correlations of hyperactivity and inattention with life satisfaction. The researchers labeled 95% confidence interval (BCa 95%CI) and calculated bias-corrected for each a*b product. An obvious mediation was indicated by the BCa 95%CI excluding 0. To avoid multicollinearity, all the continuous variables were coped with standards before the regression analyses.

## Results

### Demographic features of the objects

In total,297 questionnaires were distributed, and 291 copies were collected. Table 1 showed the demographic features of the issues and the allocations of life satisfaction. There were 120 male subjects and 171 subjects

**Table 1.** Demographic features of the objects (*N=291*)

| Variables | Category | Number | Percentage | life satisfaction (Mean±SD) |
| --- | --- | --- | --- | --- |
| Gender | Male | 120 | 41.2 | 24.46±5.97 |
|  | Female | 171 | 58.8 | 24.02±5.62 |
| Age group |  |  |  |  |
|  | 17-20 | 196 | 67.4 | 24.17±5.64 |
|  | 21-23 | 95 | 32.6 | 24.25±6.04 |
| Race |  |  |  |  |
|  | Han | 251 | 86.3 | 24.35±5.76 |
|  | Hui | 6 | 2.1 | 22.17±4.07 |
|  | Man | 25 | 8.6 | 23.92±5.98 |
|  | Other | 9 | 3.0 | 22.22±6.24 |
| Place of residence |  |  |  |  |
|  | Cities | 192 | 66.0 | 24.96±6.12 |
|  | Towns | 59 | 20.3 | 22.81±4.57 |
|  | Villages | 40 | 13.7 | 22.58±4.89 |
| Academic year |  |  |  |  |
|  | First year | 72 | 27.7 | 25.88±6.24 |
|  | Second year | 75 | 25.8 | 23.75±5.01 |
|  | Third year | 69 | 23.7 | 23.04±5.43 |
|  | Fourth year | 75 | 25.8 | 24.11±6.04 |
| The only child |  |  |  |  |
|  | Yes | 184 | 63.2 | 24.65±5.74 |
|  | No | 107 | 36.8 | 23.43±5.75 |
| Paternal educational level |  |  |  |  |
|  | Elementary education | 20 | 6.9 | 22.40±7.96 |
|  | Secondary education | 106 | 36.4 | 24.52±5.43 |
|  | Higher education | 165 | 56.7 | 24.21±5.66 |
| Maternal education level |  |  |  |  |
|  | Elementary education | 32 | 11.0 | 22.31±6.61 |
|  | Secondary education | 106 | 36.4 | 25.34±5.86 |
|  | Higher education | 153 | 52.6 | 23.80±5.38 |

Life satisfaction: SWLS

### Features of the objects between the attention deficit hyperactivity disorder (ADHD) symptomatic set and asymptomatic set

In WURS, 91 students scored ≥46 among 291 subjects. Then, the 91 people were statistically analyzed by ASRS scale, and the screening method was as follows: if the scores of 1-3 questions in Part A (namely the first six questions) were ≥3, and the scores of 4-5 questions were ≥4, and more than four of the six questions met the above standards. A total of 20 people correspond to the above conditions. These 20 subjects were divided into the ADHD symptomatic set. Table 2 presented the features of the students in the ADHD symptomatic and asymptomatic set. The female set had a lower percentage of ADHD symptoms than the male set (p = 0.025). Observe distinctions have existed between the three sets in Paternal educational level (p = 0.046). Observe distinctions weren’t existed between the two sets in age (p = 0.794), Race (p = 0.216), Place of residence (p = 0.830), Academic year (p = 0.652), The only child (p = 0.865), and Maternal education level (p = 0.520). The asymptomatic set had remarkably higher ranks of life satisfaction (p < 0.05) and resilience (p < 0.05) than the symptomatic set

**Table 2.** Features of the objects between ADHD symptomatic set and asymptomatic set (*N=291*)

| Variables | Category | ADHD | ADHD | P |
| --- | --- | --- | --- | --- |
|  |  | symptomatic | asymptomatic |  |
|  |  | N (%) | N (%) |  |
| Gender | Male | 13(10.8) | 107(89.2) | 0.025 |
|  | Female | 7(4.1) | 164(95.9) |  |
| Age group |  |  |  |  |
|  | 17-20 | 14(7.1) | 182(92.9) | 0.794 |
|  | 21-23 | 6(6.3) | 89(93.7) |  |
| Race |  |  |  |  |
|  | Han | 16(6.4) | 235(93.6) | 0.216 |
|  | Hui | 0 | 6(100.0) |  |
|  | Man | 4(16.0) | 21(84.0) |  |
|  | Other | 0 | 9(100.0) |  |
| Place of residence |  |  |  |  |
|  | Cities | 14(7.3) | 178(92.7) | 0.830 |
|  | Towns | 3(5.1) | 56(94.9) |  |
|  | Villages | 3(7.5) | 37(92.5) |  |
| Academic year |  |  |  |  |
|  | First year | 4(5.6) | 68(94.4) | 0.652 |
|  | Second year | 4(5.3) | 71(94.7) |  |
|  | Third year | 7(10.1) | 62(89.9) |  |
|  | Fourth year | 5(6.7) | 70(93.3) |  |
| The only child |  |  |  |  |
|  | Yes | 13(7.1) | 171(92.9) | 0.865 |
|  | No | 7(6.5) | 100(93.5) |  |
| Paternal educational level |  |  |  |  |
|  | Primary school and below | 4(20.0) | 16(80.0) | 0.046 |
|  | Secondary school | 5(4.7) | 101(95.3) |  |
|  | College and above | 11(6.7) | 154(93.3) |  |
| Maternal education level |  |  |  |  |
|  | Primary school and below | 3(9.4) | 29(90.6) | 0.520 |
|  | Secondary school | 5(4.7) | 101(95.3) |  |
|  | College and above | 12(7.8) | 141(92.2) |  |
| | | Mean ± SD | Mean ± SD | |
| Life Satisfaction | | 25.75±6.94 | 24.08±5.67 | 0.045 |
| Resilience | | 70.85±18.57 | 66.45±17.21 | 0.005 |

Life satisfaction: SWLS; Resilience :CD-RISC;

### Pearson correlated results in Life Satisfaction, resilience and ADHD symptoms

Table 3 demonstrated the means, standard deviations (SD), and correlated results in life satisfaction, resilience, inattention and hyperactivity. The table indicated that both hyperactivity and inattention were negatively linked to life satisfaction (hyperactivity: r = − 0.190, p < 0.05; inattention: r = − 0.121, p < 0.05) and resilience (hyperactivity: r = − 0.258, p < 0.05; inattention: r = − 0.177, p < 0.05). Life satisfaction was observably positively correlated with resilience (r = 0.752, p < 0.05).

**Table 3.** Correlated results in Life Satisfaction, resilience and ADHD symptoms.

| Variables | Mean | SD | 1 | 2 | 3 | 4 |
| --- | --- | --- | --- | --- | --- | --- |
| 1. Life Satisfaction | 24.20 | 5.77 | 1 |  |  |  |
| 2. Resilience | 66.75 | 17.31 | 0.752** | 1 |  |  |
| 3. Hyperactivity | 11.78 | 7.03 | -0.190** | -0.258** | 1 |  |
| 4. Inattention | 15.22 | 6.44 | -0.121** | -0.177** | 0.800** | 1 |

\*\*: $P < 0.05$ (two-tailed )

### The consequences of Hierarchical linear regression

Table 4 indicated the consequences of the hierarchical regression concerning life satisfaction. Hyperactivity and inattention explained 4% of the differences in life satisfaction. It was revealed that a significant negative correlation between hyperactivity and life satisfaction after controlling age and gender, (β = -0.26, p < 0.05). A quite positive consequence of resilience was indicated in life satisfaction. (β = 0.75, p < 0.05), explaining 53% of the variable of life satisfaction

**Table 4.** The results of Hierarchical linear regression analyses.

| Variables | Step 1 $\beta(95\%CI)$ | Step 2 $\beta(95\%CI)$ | Step 3 $\beta(95\%CI)$ |
| --- | --- | --- | --- |
| Step 1 |  |  |  |
| Age | -0.42(-0.68~0.32) | -0.03(-0.65~0.33) | -0.01(-0.40~0.25) |
| Gender | -0.04(-1.89~0.85) | -0.55(-1.97~0.73) | -0.06(-1.69~0.12) |
| Step 2 |  |  |  |
| inattention |  | 0.08(-0.09~0.25) | 0.02(-0.08~0.14) |
| Hyperactivity |  | -0.26**(-0.37~-0.05) | -0.02(-0.12~0.08) |
| Step 3 |  |  |  |
| Resilience |  |  | 0.75**(0.22-0.27) |
| F | 0.457 | 3.163** | 75.61** |
| R<sup>2</sup> | 0.003 | 0.042 | 0.570 |
| $\Delta R^2$ | 0.003 | 0.039 | 0.528 |

\*\*: $P < 0.05$ ( two-tailed)

### The mediation of resilience in the correlations between life satisfaction and ADHD symptoms

Table 5 revealed the magnitude of the mediation (a*b), BCa 95%CI and the path coefficients. Resilience has a significant mediating effect between life satisfaction and inattention of students. (a*b=-0.199,P<0.05) Resilience significantly moderated the correlation between life satisfaction and hyperactivity in the subjects.(a*b=-0.159,P<0.05).

**Table 5.** Mediation of resilience in the correlation between Life Satisfaction and ADHD symptoms.

| ADHD symptoms |  |  |  |  |  |
| --- | --- | --- | --- | --- | --- |
| Predictor | Path coefficient | | | | $a*b$(Bca 95%CI) |
| Variables |  |  |  |  |  |
|  | c | a | b | C' |  |
| inattention | -0.108** | -0.476** | 0.251** | 0.011 | -0.199**(-0.028~-0.002) |
| Hyperactivity | -0.156** | -0.635** | 0.251** | 0.003 | -0.159**(-0.266~-0.057) |

\*\*: $P < 0.05$ ;
c: correlations between life satisfaction and ADHD symptoms;
a: correlations between resilience and ADHD symptoms;
b: correlations between life satisfaction and resilience after controlling the predictors;
c': correlations between life satisfaction and ADHD symptoms after using resilience as the mediating variable;

## Discussion

UP till now, there has been little investigation into ADHD among dental students. Existing studies have explored the awareness and knowledge of dental students about ADHD but have not focused on ADHD in the group itself [32]. This study focuses on ADHD symptoms among dental students. It is also the first to explore the correlation between life satisfaction and ADHD symptoms of dental students, as well as to verify the mediation of mental resilience in these relationships. On one hand, the outcomes of this study indicate that 6.87% of dental students have symptomatic ADHD. The relevant data of Chinese dental students was lower than the morbidity rate of ADHD (8.45%) among Chinese medical students [33]. Several studies have demonstrated a correlation between ADHD and learning disabilities, particularly in verbal reasoning, visual-auditory memory, and cognitive traits [34,35]. It can be inferred that some individuals with ADHD will be slightly worse than ordinary people in learning ability. Chinese undergraduates must take a national exam before being admitted and choose their prospective colleges and majors based on their scores. In most Chinese medical universities, the average score of students admitted to the stomatology program is higher than that of undergraduates. majoring in clinical medicine in the same institution. Therefore, among high school graduates with ADHD symptoms who want to apply for stomatology, compared with graduates who want to use it for clinical medicine, most of them cannot study oral medication due to the entrance examination scores. On the other hand, based on the data from WHO, the result is higher than the worldwide prevalence of ADHD (2.8%) among adults [5]. This data may indicate that, compared with the mental disorders which are easily recognized and detected, dental students facing heavy academic and life pressure often underestimate or ignore ADHD symptoms. Early detection and diagnosis of dental students with ADHD symptoms are necessary to treat them as early as possible. The research reveals that the morbidity of ADHD significantly differs in students whose fathers have different levels of education. The probable reason is that Chinese fathers teach their children more harshly, while fathers with less education often cannot control their emotions and often criticize their children. These may make a difference on the recovery of ADHD symptoms in nonage. This suggests that the relevant department should improve the education level of residents. A review of three decades of research shows that, in the long run, people with untreated ADHD are impaired in social functioning, self-esteem, and occupation compared to ordinary people [36]. The resilience level of the symptomatic set is observably lower than that of the asymptomatic group, which is consistent with the study of medical students [33]. The rational interpretation is that people with ADHD often have other mental disorders, including mood disorders, learning disabilities, and so on [34,37], and many have experienced significant adverse events [38]. These are all inversely related to resilience [33].

One of the principal findings of this research was that life satisfaction had significant correlations with hyperactivity among people who major in dentistry. In contrast, there was no correlation between life satisfaction and inattention after controlling the distracting factors. This is different from existing research. In the past literature, some show that inattention is correlated with life satisfaction[33], while others believe that there is no apparent difference between diverse subtypes of ADHD[39]. There are two reasons for this result: First, ADHD is more general in men than women, and it is more common for men to show hyperactivity than inattention. Second, the survey uses recall scales. Hyperactivity is more likely to attract the attention of others, such as teachers and parents. These individuals mentioned hyperactivity around subjects more often than inattention, which may make issues memorize hyperactivity more clearly than inattention.

Another important finding of the research is that resilience significantly mediates between life satisfaction and the two ADHD symptoms in people who major in dentistry. Besides being directly related to satisfaction in life, the two ADHD symptoms are also indirectly associated with life satisfaction through the mediation of resilience. Students with more obvious ADHD symptoms had worse resilient standards. It was associated with worse life satisfaction in turn. And vice versa. Unlike traditional psychology, which focuses on negative and pathological psychology, positive psychology uses human motivation as its basic tenet. Since positive psychology was proposed, it has been increasingly widely used to prevent and treat psychological disorders in the past 20 years. Resilience is an essential component of positive psychology, which refers to the excellent adaptability of individuals encountering adversity, scar and other remarkable pressures in life [40]. A meta-analysis in different populations showed that multiple training programs and interventions had a specific effect [41]. Previous studies have shown that appropriate interventions to improve resilience are feasible among university faculty and military personnel [42,43]. Interventions based on mindfulness and cognitive-behavioural have been used more frequently in the psychological treatment of ADHD. This suggests that such interventions are effective in improving

ADHD symptoms in adults [44,45]. Therefore, on an evidence-based basis, the School of Stomatology can design a targeted resilience intervention program to enhance the satisfaction in the life of students with ADHD. It can improve the enthusiasm of these students for study and life, which is conducive to cultivating more excellent future dentists. The consequences of this research explained 57% of the differences in students’ life satisfaction. The model didn’t cover some potential variables, such as other social and psychological factors, which may affect students’ satisfaction with life.

This study chose dental students as the subject group and obtained a large sample size and a highly effective response rate. This paper explored the correlation between life satisfaction and the two sub-symptoms of ADHD and analyzed the related mechanism between the two sub-symptoms and resilience. Admittedly, the research exists some boundedness. First, the studies all used self-report questionnaires, and data collected on these questionnaires can introduce errors. However, self-report questionnaires are often used in psychiatric clinical practice and large-scale epidemiological studies [46]. Second, as a cross-sectional investigation, it is incapable of exploring the causal relationship between the variables, and it is difficult to follow up on the subjects. Relevant prospective cohort studies can be carried out to supplement it. Third, limited by resource allocation, multi-centre research among multi-national, multi-regional, and multi-level medical schools should be carried out in the future.

## Conclusions

Studies have shown that the morbidity of ADHD in general adults is lower than Chinese dental students. It was revealed that paternal education level was relevant to the morbidity. Of the two ADHD symptoms, only hyperactivity was significantly associated with life satisfaction. Distinct mediation of resilience was found between life satisfaction and ADHD symptoms. The consequences of the study can ensure the early detection of ADHD symptoms in dental students to enhance the mental health level of future dentists to a certain extent. Resilience intervention programs can improve dental students’ life satisfaction, especially for students with ADHD symptoms.

## Data Availability

All data files are available from the Dryad database (accession number https://doi.org/10.5061/dryad.51c59zwd0).

## Acknowledgements

The authors give expressions of gratitude to all the students participating in this experiment and the faculty of stomatology who helped to collect the data.

## Author Contributions

Conceptualization, Y.S. and C.T.; methodology, Y.S., L.M. and C.T.; formal analysis, Y.S.; investigation, Y.S., L.M., S.C. and Z.P.; resources, L.M. and C.T.; data curation, Y.S.; writing—original draft preparation, Y.S.; writing—review and editing, Y.S., S.C., Z.P. and C.T.; visualization, Y.S. and C.T.; supervision, C.T.; project administration, L.M. and C.T. All authors have read and agreed to the published version of the manuscript.

## Institutional Review Board Statement

The study was conducted in accordance with the Declaration of Helsinki, and approved by the Ethics Committee of China Medical University on May 12th with protocol code 2023129.

## Informed Consent Statement

Informed consent was obtained from all subjects involved in the study.

## Data Availability Statement

Publicly available datasets were analyzed in this study. This data can be found here: https://doi.org/10.5061/dryad.51c59zwd0

## References

1. GBD 2019 Mental Disorders Collaborators. Global, regional, and national burden of 12 mental disorders in 204 countries and territories, 1990-2019: a systematic analysis for the Global Burden of Disease Study 2019. Lancet Psychiatry. 2022 Feb;9(2):137–50.

2. Chhibber A, Watanabe AH, Chaisai C, Veettil SK, Chaiyakunapruk N. Global Economic Burden of Attention-Deficit/Hyperactivity Disorder: A Systematic Review. PharmacoEconomics. 2021 Apr;39(4):399–420.

3. Association AP. Diagnostic and Statistical Manual of Mental Disorders, 5th Edition: DSM-5. 5th edition. Washington, D.C: American Psychiatric Publishing; 2013. 991 p.

4. Association AP, Kennedy PJ. Understanding Mental Disorders: Your Guide to DSM-5. 1st edition. Washington, DC: American Psychiatric Pubublishing; 2015. 388 p.

5. Fayyad J, Sampson NA, Hwang I, Adamowski T, Aguilar-Gaxiola S, Al-Hamzawi A, et al. The descriptive epidemiology of DSM-IV Adult ADHD in the World Health Organization World Mental Health Surveys. Atten Deficit Hyperact Disord. 2017 Mar;9(1):47–65.

6. Harpin VA. The effect of ADHD on the life of an individual, their family, and community from preschool to adult life. Arch Dis Child. 2005 Feb;90 Suppl 1(Suppl 1):i2–7.

7. Schwanz KA, Palm LJ, Brallier SA. Attention problems and hyperactivity as predictors of college grade point average. J Atten Disord. 2007 Nov;11(3):368–73.

8. Murphy K. Psychosocial treatments for ADHD in teens and adults: a practice-friendly review. J Clin Psychol. 2005 May;61(5):607–19.

9. Roberts W, Peters JR, Adams ZW, Lynam DR, Milich R. Identifying the facets of impulsivity that explain the relation between ADHD symptoms and substance use in a nonclinical sample. Addict Behav. 2014 Aug;39(8):1272–7.

10. Tong L, Shi HJ, Zhang Z, Yuan Y, Xia ZJ, Jiang XX, et al. Mediating effect of anxiety and depression on the relationship between Attention-deficit/hyperactivity disorder symptoms and smoking/drinking. Sci Rep. 2016 Feb 29;6:21609.

11. Shen Y, Zhang Y, Chan BSM, Meng F, Yang T, Luo X, et al. Association of ADHD symptoms, depression and suicidal behaviors with anxiety in Chinese medical college students. BMC Psychiatry.2020 Apr 22;20(1):180.

12. Shen Y, Chan BSM, Huang C, Cui X, Liu J, Lu J, et al. Suicidal behaviors and attention deficit hyperactivity disorder (ADHD): a cross-sectional study among Chinese medical college students. BMC Psychiatry. 2021 May 18;21(1):258.

13. Shi M, Du TJ. Associations of personality traits with internet addiction in Chinese medical students: the mediating role of attention-deficit/hyperactivity disorder symptoms. BMC Psychiatry. 2019 Jun 17;19(1):183.

14. Hyman SE. The diagnosis of mental disorders: the problem of reification. Annu Rev Clin Psychol. 2010;6:155–79.

15. Das D, Cherbuin N, Butterworth P, Anstey KJ, Easteal S. A Population-Based Study of Attention Deficit/Hyperactivity Disorder Symptoms and Associated Impairment in Middle-Aged Adults. PLoS ONE. 2012 Feb 8;7(2):e31500.

16. Benoit V, Gabola P. Effects of Positive Psychology Interventions on the Well-Being of Young Children: A Systematic Literature Review. Int J Environ Res Public Health. 2021 Nov 17;18(22):12065.

17. Bazargan-Hejazi S, Shirazi A, Wang A, Shlobin NA, Karunungan K, Shulman J, et al. Contribution of a positive psychology-based conceptual framework in reducing physician burnout and improving well-being: a systematic review. BMC Med Educ. 2021 Nov 25;21(1):593.

18. Jarrett T. Warrior Resilience Training in Operation Iraqi Freedom: combining rational emotive behavior therapy, resiliency, and positive psychology. US Army Med Dep J. 2008;32–8.

19. Greven CU, Buitelaar JK, Salum GA. From positive psychology to psychopathology: the continuum of attention-deficit hyperactivity disorder. J Child Psychol Psychiatry. 2018 Mar;59(3):203–12.

20. Sedgwick JA, Merwood A, Asherson P. The positive aspects of attention deficit hyperactivity disorder: a qualitative investigation of successful adults with ADHD. Atten Deficit Hyperact Disord. 2019 Sep;11(3):241–53.

21. Carr A. Positive Psychology: The Science of Happiness and Human Strengths. Psychology Press; 2004. 412 p.

22. Spaccarelli S. Stress, appraisal, and coping in child sexual abuse: a theoretical and empirical review. Psychol Bull. 1994 Sep;116(2):340–62.

23. Yeh CB, Gau SSF, Kessler RC, Wu YY. Psychometric properties of the Chinese version of the adult ADHD Self-report Scale. Int J Methods Psychiatr Res. 2008;17(1):45–54.

24. Kessler RC, Ustün TB. The World Mental Health (WMH) Survey Initiative Version of the World Health Organization (WHO) Composite International Diagnostic Interview (CIDI). Int J Methods Psychiatr Res. 2004;13(2):93–121.

25. Ni HC, Gau SSF. Co-occurrence of attention-deficit hyperactivity disorder symptoms with other psychopathology in young adults: parenting style as a moderator. Compr Psychiatry. 2015 Feb 1;57:85–96.

26. Ward MF, Wender PH, Reimherr FW. The Wender Utah Rating Scale: an aid in the retrospective diagnosis of childhood attention deficit hyperactivity disorder. Am J Psychiatry. 1993 Jun;150(6):885–90.

27. Diener E, Emmons RA, Larsen RJ, Griffin S. The Satisfaction With Life Scale. J Pers Assess. 1985 Feb;49(1):71–5.

28. Shi M, Wang X, Bian Y, Wang L. The mediating role of resilience in the relationship between stress and life satisfaction among Chinese medical students: a cross-sectional study. BMC Med Educ. 2015 Feb 13;15(1):16.

29. Pavot W, Diener E, Colvin R, Sandvik E. Further Validation of the Satisfaction With Life Scale: Evidence for the Cross-Method Convergence of Well-Being Measures. J Pers Assess. 1991 Sep 1;57:149–61.

30. Connor K, Davidson J. Development of a new resilience scale: The Connor-Davidson Resilience Scale (CD-RISC). Depress Anxiety. 2003 Sep 1;18:76–82.

31. Windle G, Bennett K, Noyes J. A methodological review of resilience measurement scales. Health Qual Life Outcomes. 2011 Jan 1;9:1–8.

32. Chopra A, Vishnupriya V, Gayathri R, Kavitha S. Knowledge and awareness of dental students on attention deficit hyperactivity disorder. J Adv Pharm Technol Res. 2022 Nov;13(Suppl 1):S308–13.

33. Shi M, Liu L, Sun X, Wang L. Associations between symptoms of attention-deficit/ hyperactivity disorder and life satisfaction in medical students: the mediating effect of resilience. BMC Med Educ. 2018 Jul 13;18:164.

34. Talero-Gutiérrez C, Velez-van-Meerbeke A, Reyes R. A Clinical Study of ADHD Symptoms With Relation to Symptoms of Learning Disorders in Schoolchildren in Bogota, Colombia. J Atten Disord. 2010 Oct 1;16:157–63.

35. Toffalini E, Buono S, Cornoldi C. The structure, profile, and diagnostic significance of intelligence in children with ADHD are impressively similar to those of children with a specific learning disorder. Res Dev Disabil. 2022 Oct 1;129:104306.

36. Shaw M, Hodgkins P, Caci H, Young S, Kahle J, Woods AG, et al. A systematic review and analysis of long-term outcomes in attention deficit hyperactivity disorder: effects of treatment and non-treatment. BMC Med. 2012 Sep 4;10:99.

37. Cumyn L, French L, Hechtman L. Comorbidity in Adults with Attention-Deficit Hyperactivity Disorder. Can J Psychiatry. 2009 Oct 1;54(10):673–83.

38. Dvorsky MR, Langberg JM. A Review of Factors that Promote Resilience in Youth with ADHD and ADHD Symptoms. Clin Child Fam Psychol Rev. 2016 Dec 1;19(4):368–91.

39. O’Brien JW, Dowell LR, Mostofsky SH, Denckla MB, Mahone EM. Neuropsychological profile of executive function in girls with attention-deficit/hyperactivity disorder. Arch Clin Neuropsychol Off J Natl Acad Neuropsychol. 2010 Nov;25(7):656–70.

40. Newman R. APA’s resilience initiative. Prof Psychol Res Pract. 2005;36:227–9.

41. Joyce S, Shand F, Tighe J, Laurent SJ, Bryant RA, Harvey SB. Road to resilience: a systematic review and meta-analysis of resilience training programmes and interventions. BMJ Open. 2018 Jun 14;8(6):e017858.

42. Johnson DC, Thom NJ, Stanley EA, Haase L, Simmons AN, Shih P an B, et al. Modifying Resilience Mechanisms in At-Risk Individuals: A Controlled Study of Mindfulness Training in Marines Preparing for Deployment. Am J Psychiatry. 2014 Aug;171(8):844–53.

43. Sood A, Prasad K, Schroeder D, Varkey P. Stress Management and Resilience Training Among Department of Medicine Faculty: A Pilot Randomized Clinical Trial. J Gen Intern Med. 2011 Aug;26(8):858–61.

44. Geffen J, Forster K. Treatment of adult ADHD: a clinical perspective. Ther Adv Psychopharmacol. 2018 Jan;8(1):25–32.

45. Kolar D, Keller A, Golfinopoulos M, Cumyn L, Syer C, Hechtman L. Treatment of adults with attention-deficit/hyperactivity disorder. Neuropsychiatr Dis Treat. 2008 Feb;4(1):107–21.

46. Magnússon P, Smári J, Sigurdardóttir D, Baldursson G, Sigmundsson J, Kristjánsson K, et al. Validity of self-report and informant rating scales of adult ADHD symptoms in comparison with a semistructured diagnostic interview. J Atten Disord. 2006 Feb;9(3):494–503.

